# In search of “hepatic factor:” Lack of evidence for ALK1 ligands BMP9 and BMP10

**DOI:** 10.1101/2020.07.09.20148320

**Authors:** Teresa L. Capasso, Sara M. Trucco, Morgan Hindes, Tristin Schwartze, Jamie L. Bloch, Jacqueline Kreutzer, Stephen C. Cook, Cynthia S. Hinck, Davide Treggiari, Brian Feingold, Andrew P. Hinck, Beth L. Roman

## Abstract

In children with single ventricle physiology, the Glenn procedure is performed to redirect venous return from the superior vena cava directly to the pulmonary arteries and route venous return from the inferior vena cava exclusively to the systemic circulation. Although this surgery successfully palliates the hemodynamic stress experienced by the single ventricle, patients frequently develop pulmonary arteriovenous malformations (PAVMs). Interestingly, PAVMs may regress upon rerouting of hepatic venous effluent to the pulmonary vasculature, suggesting the presence of a circulating “hepatic factor” that is required to prevent PAVMs. Here, we test the hypothesis that hepatic factor is bone morphogenetic protein 9 (BMP9) and/or BMP10. These circulating ligands are produced by the liver and activate endothelial endoglin (ENG)/ALK1 signaling, and mutations in *ENG* and *ALK1* cause hereditary hemorrhagic telangiectasia, a genetic disease associated with AVM development. However, we found no within-subject variation in BMP9, BMP10, or BMP9/10 plasma concentrations when sampled from five cardiovascular sites, failing to support the idea that the Glenn would limit access of these ligands to the lung vasculature. Unexpectedly, however, we found a significant decrease in all three ligand concentrations in Glenn cases versus controls. Our findings suggest that BMP9/BMP10/ENG/ALK1 signaling may be decreased in the Glenn vasculature but fail to implicate these ligands as hepatic factor.

## Introduction

Considerable evidence suggests that the liver produces or modifies a circulating factor critical for preventing pulmonary arteriovenous malformations (PAVMs). In hepatopulmonary syndrome (HPS), liver dysfunction is associated with hypoxemia secondary to PAVMs, and PAVMs are reversed by liver transplantation (1). Additionally, portosystemic shunts that allow gut venous effluent to bypass the liver lead to PAVMs, which resolve when the shunt is closed (2). Further evidence comes from single-ventricle patients who undergo a three-staged surgery to relieve hemodynamic burden on the heart and correct oxygen desaturation. The second-stage surgery, the bidirectional Glenn, directs passively draining venous return from the superior vena cava (SVC) to the pulmonary circulation, with venous return from the inferior vena cava (IVC) pumped to the systemic circulation. Although the Glenn effectively decreases ventricular hemodynamic stress, intrapulmonary arteriovenous shunting is pervasive, and up to 25% develop clinically significant hypoxemia secondary to diffuse PAVMs (3, 4). While early theories of PAVM development focused on the absence of pulsatile flow or increased lower lobe perfusion (3), later evidence implicated the exclusion of a liver-derived substance from the pulmonary vasculature. This “hepatic factor” was postulated based on correlation between laterality of PAVMs and laterality of exclusion of hepatic venous effluent (5), and its existence is strongly supported by evidence that the third-stage Fontan procedure (completion of the total cavopulmonary anastomosis), which reroutes IVC flow to the lungs without restoring pulsatility, is strongly associated with PAVM regression (6). Despite the strong evidence for hepatic factor, its identity remains unknown.

Approximately 80% of PAVMs are associated with hereditary hemorrhagic telangiectasia (HHT), a genetic disorder caused primarily by mutations in bone morphogenetic protein (BMP) receptors endoglin (*ENG*) and ALK1 (encoded by *ACVRL1*) (7). This pathway is active in lung endothelium and ligands include BMP9 and BMP10 homodimers and BMP9/10 heterodimer (8, 9). Both *BMP9* and *BMP10* are transcribed in hepatic stellate cells (9). Given the strong relationship between PAVMs and HHT, the hepatic origins of *BMP9* and *BMP10*, and evidence of decreased plasma BMP9 in HPS (10), we hypothesized that ALK1 ligands may be the hepatic factor required for PAVM prevention. We expect that hepatic factor is either labile or actively removed from circulation on first pass through the systemic circulation, making it unavailable to the lung vasculature in Glenn circulation. Accordingly, in normal circulation, we hypothesized that concentrations of ALK1 ligands would be higher in the right atrium and pulmonary artery compared to the SVC and infrahepatic IVC.

## Methods

This study was approved by the University of Pittsburgh Institutional Review Board. Participants undergoing clinically-indicated cardiac catheterization were recruited between September 2015 and February 2017 and provided informed child assent and/or parental consent. Patients with bidirectional Glenn, prior to Fontan, were compared to two-ventricle controls. Excluded diagnoses among controls included single ventricle physiology, unrepaired complex congenital heart disease, and large shunt lesions. Patients with liver disease, anemia (hemoglobin < 8 g/dL), cardiac surgery within 30 days, or transfusion within 48 hours were excluded from both cohorts.

We collected 1 ml blood in K_2_EDTA tubes from five sites: the right atrium, pulmonary artery, aorta, SVC, and infrahepatic IVC. We measured ligands in duplicate in 30 μL of plasma via sandwich ELISAs (R&D Systems, Minneapolis, USA) using DY3209 (BMP9), MAB2926 and BAF3956 (BMP10), and MAB2926 and BAF3209 (BMP9/10), using in-house generated recombinant proteins for the latter two standard curves. We fit data to a four parameter logistic curve and performed statistical analysis using GraphPad Prism (San Diego, USA). We ran all samples from an individual on a single plate, and the operator was blinded to sample identity. Sample volume limitations prevented us from assaying all ligands in every individual.

## Results

Diagnoses in 38 controls [mean age 5.8 years (4 months - 12.6 years); 21 males, 17 females] included: small shunt lesions (21), repaired forms of congenital heart disease with two-ventricle physiology (11), vascular stenosis (5), valvar obstructive lesions (2), and hypertrophic cardiomyopathy (1). Primary cardiac diagnoses in 9 Glenn cases [mean age 2.9 years (range 22 months to 5.1 years); 7 males, 2 females] included: variants of hypoplastic left heart syndrome (5); pulmonary atresia with intact ventricular septum (2); double outlet right ventricle with pulmonary atresia (1); and heterotaxy with right atrial isomerism (1).

BMPs are generated as proprotein dimers that are cleaved between the N-terminal prodomains and C-terminal growth factor (GF) domains, releasing the disulfide-bonded GF dimer (GFD). In control plasma, we detected BMP9 GFD, BMP10 proprotein, and BMP9/10 GFD (Fig. 1), but not BMP10 GFD (DY2926, R&D Systems; data not shown). However, we found no differences in plasma concentrations of any ALK1 ligand when comparing within-subject values across the right atrium, pulmonary artery, aorta, SVC, and IVC (Fig. 1). This result suggests that these ligands are neither particularly labile nor actively removed on first pass through the systemic or pulmonary circulation, failing to support the hypothesis that they represent the hepatic factor required to prevent PAVMs.

**Figure 1.**
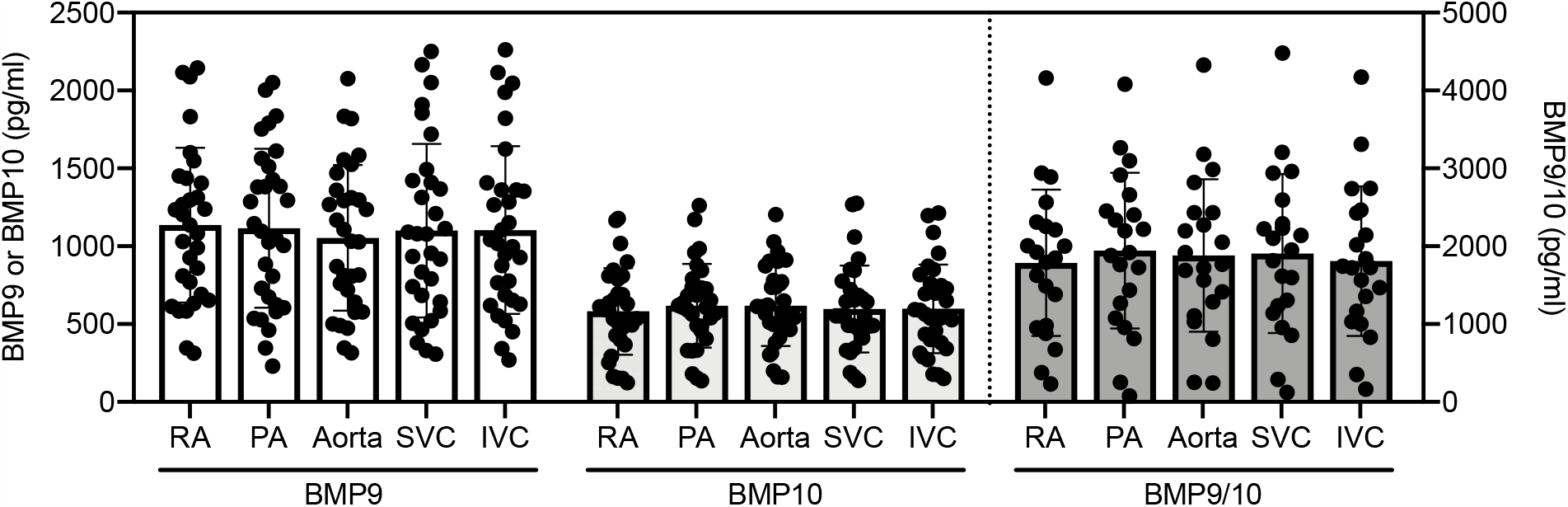
Plasma BMP9, BMP10, and BMP9/10 concentrations do not differ between pulmonary inflow and systemic venous circulation. Plasma from controls was sampled from the right atrium (RA), pulmonary artery (PA), aorta, SVC, and infrahepatic IVC and ligands quantified by sandwich ELISA. All values are GFD equivalents. Error bars, mean ± SD. Not significant by repeated measures one-way ANOVA: BMP9, N = 31, *P*=0.13; BMP10, N=31, *P*=0.17; BMP9/10, N=21, *P*=0.21.

Evaluation of plasma from Glenn cases similarly revealed no significant differences in within-subject ligand concentrations across sampling sites (data not shown). However, after collapsing data across sites, comparison of grand means revealed significant decreases in plasma concentrations of all ligands in Glenn cases compared to controls (Fig. 2), and significance persisted after age-adjustment (multiple linear regression; BMP9, *P* = 0.04; BMP10, *P* = 0.0002, BMP9/10, *P* = 0.002). Although our sample set is underpowered, we saw no correlation between ligand concentration and PAVMs (Fig. 2).

**Figure 2.**
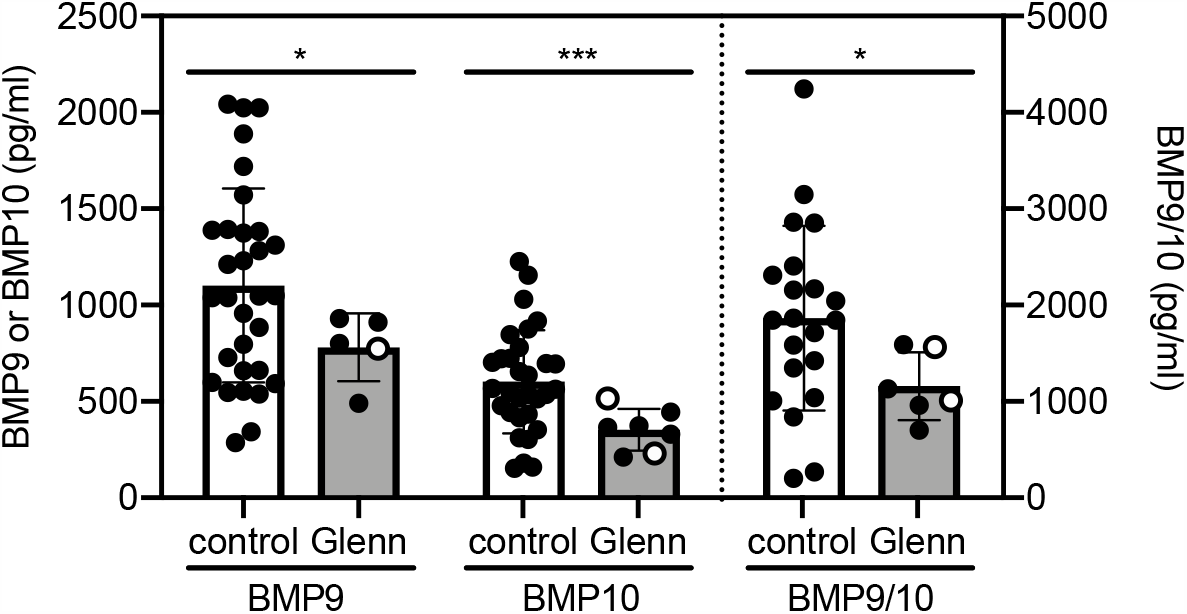
Plasma BMP9, BMP10, and BMP9/10 concentrations are lower in Glenn cases compared to controls. Within-patient data were averaged across all sampling sites (RA, PA, aorta, SVC, IVC) for controls and Glenn cases and evaluated by Welch’s *t*-test. Error bars, mean ± SD. BMP9: control, N=31; Glenn, N=5; *P=*0.02. BMP10: control, N=31; Glenn, N=7; *P*=0.0006. BMP9/10: control, N=21; Glenn, N=6; *P*=0.01. Open circles indicate cases with PAVMs.

## Discussion

We found no within-subject differences in plasma concentrations of BMP9 GFD, BMP10 proprotein, or BMP9/10 GFD across different sampling sites, in agreement with a recent report regarding BMP9 GFD (11) and failing to support the idea that ALK1 ligands are the “hepatic factor” required to prevent PAVMs. However, it remains possible that liver-derived BMP9 or BMP9/10 proproteins (not assayed) may exhibit site-dependent concentration differences, or that enzymes required to cleave proproteins are unavailable in the Glenn circulation. Surprisingly, we found that Glenn cases had significantly lower concentrations of all three ligands compared to controls. Measurement of these ligands in additional Glenn cases and in Fontan cases will be required to determine the biological significance of this finding with respect to Glenn-associated PAVMs.

## Data Availability

All data are archived on secure servers maintained by the University of Pittsburgh Department of Human Genetics

